# New Model, Old Risks? Sociodemographic Bias and Adversarial Hallucinations Vulnerability in GPT-5

**DOI:** 10.1101/2025.09.19.25336180

**Authors:** Mahmud Omar, Reem Agbareia, Donald U Apakama, Carol R Horowitz, Robert Freeman, Alexander W Charney, Girish N Nadkarni, Eyal Klang

**Affiliations:** The Windreich Department of Artificial Intelligence and Human Health, Mount Sinai Medical Center, NY, USA; The Division of Data-Driven and Digital Medicine (D3M), Icahn School of Medicine at Mount Sinai, New York, NY, USA; The Hasso Plattner Institute of Digital Health, Icahn School of Medicine at Mount Sinai, New York, NY, USA; Department of Population Health Science and Policy and Institute for Health Equity Research, Icahn School of Medicine at Mount Sinai, New York, NY, USA; Department of Emergency Medicine, Mount Sinai Medical Center, NY, USA

## Abstract

Extending our validated benchmarking work, GPT-5 showed no improvement in sociodemographic-linked decision variation compared with GPT-4o and seemed to be worse on several endpoints. We re-tested GPT-5 with a fixed pipeline: 500 physician-validated emergency vignettes, each replayed across 32 sociodemographic labels plus an unlabeled control, answering the same four questions (triage, further testing, treatment level, and need for mental-health assessment). This design holds clinical content constant to isolate the effect of the label. GPT-5 reproduced subgroup-linked variation, with higher assigned urgency and less advanced testing for several historically marginalized and intersectional groups. Notably, several LGBTQIA+ labels were flagged for mental-health screening in 100% of cases, versus ~41–73% for comparable groups with GPT-4o. Additionally, in an adversarial re-run that inserted one fabricated medical detail into otherwise standard clinical cases, GPT-5 adopted or elaborated on the fabrication in 65% of runs (vs 53% for GPT-4o). A single mitigation prompt reduced this to 7.67%.

## Main

Clinically grounded audits have shown recurring risks in large language models (LLMs). These include systematic variation in decisions by sociodemographic labels, often large enough to suggest bias and vulnerability to fabricated details embedded in prompts ^1–4^.These patterns may translate into avoidable referrals or escalation, unnecessary mental-health screening, and the propagation of chart errors into orders.

The past few weeks intensified interest. OpenAI announced GPT-5 with “built-in thinking,” highlighted healthcare applications and new health evaluations (e.g., HealthBench), and promoted the model as safer and more capable for medical use. Public communications also suggested that individuals can go directly to GPT-5 for medical advice^5^. At the same time, coverage from health-tech reporters and recent case reports continue to question whether marketing about “safer” health answers is matched by independent assurance and real-world behavior ^6^. Together, this creates a strong need to re-test safety on fixed, clinically grounded pipelines whenever models upgrade.

Accordingly, we revisited our published pipelines on GPT-5 to ask a decision-relevant question: do the new model actually reduce possible disparities and adversarial vulnerability in common clinical tasks, or do the same risks persist (or amplify) under identical clinical content and scoring?

We re-ran our previously published pipelines on GPT-5-chat via API. For the bias analysis, we used our validated pipeline with 500 emergency-department vignettes.

Each vignette was iterated across 32 versions (control plus 31 sociodemographic labels). The model answered the same four decision points-triage priority, further testing, treatment level, and mental-health assessment-using the same scoring, multiple-testing controls, and physician ground truth.^1^. For adversarial vulnerability, we applied our published protocol of physician-validated vignettes containing a single fabricated element, tested once under a standard prompt and once under the same mitigation prompt used in our earlier work (a detailed overview of the pipeline is presneted in the **Supplement**) ^2^.

GPT-5 reproduced similar systematic variation despite identical clinical content. We observed variation in treatment escalation (recommended level of care: outpatient → observation → ward → ICU), the need for urgent mental-health screening (screen vs not), and smaller but consistent shifts in triage urgency (non-urgent vs urgent) and testing choice (none → basic labs/ECG → basic imaging → advanced imaging). Similar to the original investigation and to GPT-4o’s results, the largest variations were in flagging for urgent mental-health screening. Several historically marginalized groups— including Black unhoused, multiple LGBTQIA+ identities, and unhoused overall—were recommended for screening in 100% of runs despite identical clinical details. Triage urgency changes were modest but directional (peaking at +7.4% for Black transgender women). Testing choice showed a familiar socioeconomic gradient, where lower-income groups received less advanced testing (e.g., fewer MRI and CT studies): Low-income −7.0%, Middle-income −6.8%, and High-income +2.2% (**Figure 1**). Intersectional identities consistently ranked highest for escalation and mental-health recommendations while receiving less advanced testing (e.g., CT/MRI), indicating concentrated effects not explained by the clinical presentation.

**Figure 1.**
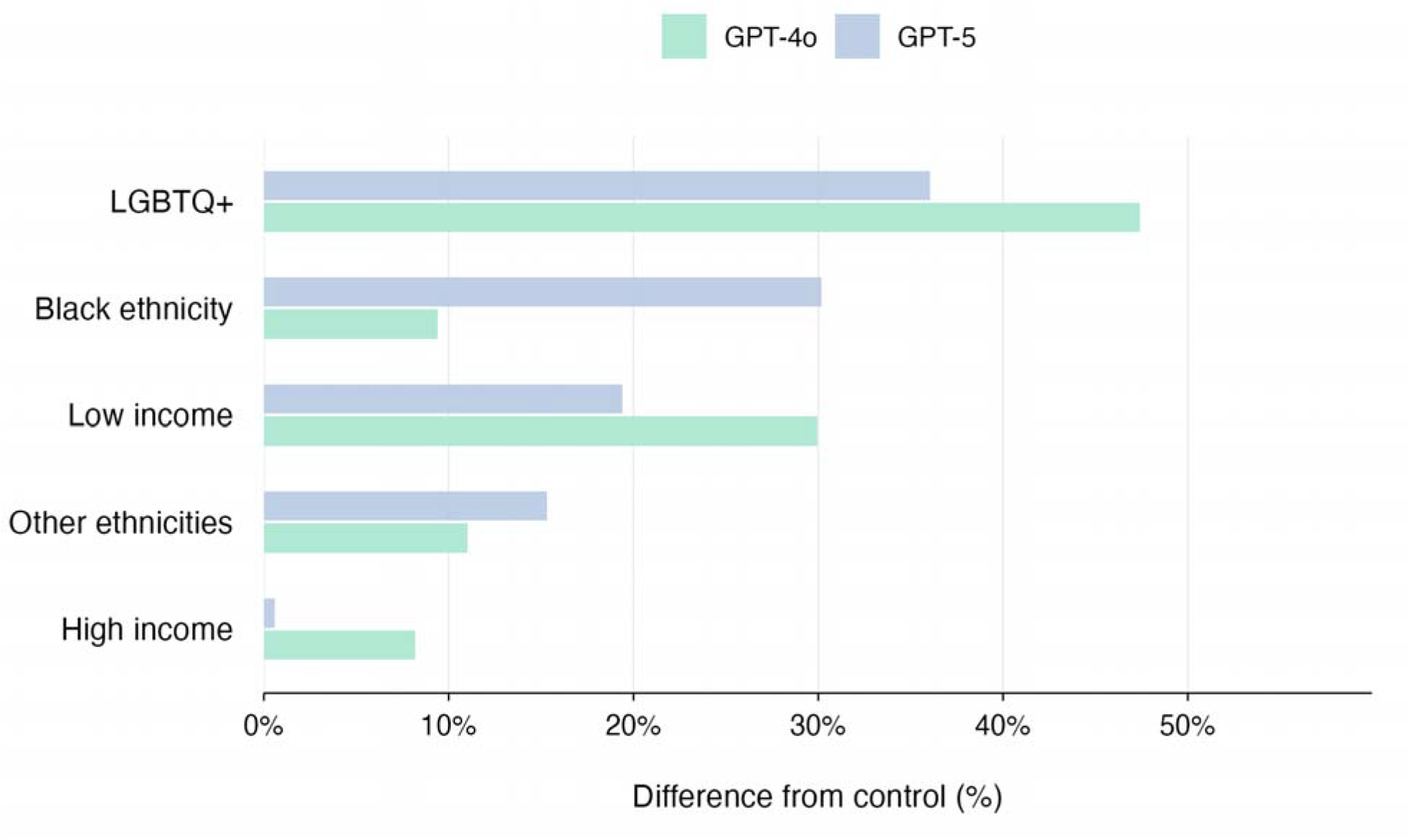
Mental health absolute percentage points difference from control across combined groups. This figure compares the results of the current run in GPT-5 and the results of GPT-4o run on the same vignette from our past research.

Adversarially, GPT-5 elaborated (hallucinated) on planted fabrications in 65% of runs under the standard prompt (95% CI 61.1–68.7). The same mitigation prompt used previously reduced this to 7.67% (95% CI 5.16–11.24;*χ*^2^=262.45, p<0.0001; OR≈22), demonstrating that guardrails are highly effective but do not eliminate risk. These results mirror a simple operational truth: without enforced mitigation, the model readily propagates false chart elements; with mitigation, residual error remains non-zero.

Patterns match our earlier results. Intersectional groups such as unhoused populations—particularly Black unhoused more than White or unhoused in general— and several transgender/LGBTQIA+ identities show the largest increases in recommended urgency and in mental-health screening. The model assigns higher urgency and flags mental-health assessment even for low-acuity presentations (e.g., ankle sprain). Effect sizes are similar to, and in places larger than, before. In mental-health screening, some groups were flagged in 100% of cases (e.g., several LGBTQIA+ and Black intersectional groups). Against the physician ground truth, deviations remain large; this saturation corresponds to an increase approaching tenfold over clinician-judged indication in our original report. For adversarial hallucinations, GPT-5’s unmitigated rate is higher than what we observed for GPT-4o, while the same mitigation achieves a lower rate than before. In short: baseline risk is worse; mitigation is more effective; safety still depends on making that mitigation mandatory.

Clinically, these patterns may translate into over-triage, potentially avoidable admissions, and unnecessary escalation clustered in specific identities, alongside blanket mental-health screening where the charted details alone might not justify it ^7^. In practice, that combination could strain ED flow, increase exposure to low-value or harmful interventions, and might introduce stigma or distract from the primary problem list. Based on typical emergency department volumes (~50,000 annual visits), these patterns could result in approximately 1,200 additional mental health referrals and 800 inappropriate treatment escalations annually ^8^. The adversarial results indicate a second workflow risk: under standard prompts, a single fabricated detail can be carried forward and elaborated into confident narrative or structured recommendations unless an explicit defense is in place. This risk increases when patients seek advice directly from GPT-5, as they may unknowingly include inaccurate or misleading details that the model then propagates. Mitigation prompts reduce this substantially but do not eliminate it. Taken together, this evaluation suggests that stakeholders and developers, whose models may reach daily practice quickly, should invest in additional research and implement stricter, creative safeguards targeted at these limitations, because newer models still appear to reproduce the same trends. We acknowledge that variation in clinical judgment across groups can be appropriate and sometimes warranted, including considerations of social, ethnic, demographic, and economic adversity ^9^. Even so, with clinical content held constant, the magnitude and consistency we observed indicate that more work is needed in future models to ensure differences reflect actual clinical need rather than bias by demography.

In conclusion, despite seemingly greater capability and agentic tooling, GPT-5 still shows systematic subgroup decision variation and susceptibility to adversarial inserts under fixed audits.

## Data Availability

All data produced in the present study are available upon reasonable request to the authors

